# Neurosurgery and Artificial Intelligence: A Metric Analysis of Scopus-Indexed Original Articles (2014-2023)

**DOI:** 10.1101/2025.04.04.25325251

**Authors:** Hector Julio Piñera-Castro, Christian Borges-García

## Abstract

**Introduction:** A comprehensive analysis of artificial intelligence’s (AI) integration into neurosurgery is vital to identify research priorities, address gaps, and inform strategies for equitable innovation.

**Objective:** To conduct a bibliometric analysis of Scopus-indexed (2014-2023) original articles at the intersection of AI and neurosurgery.

**Method:** A descriptive metric study was conducted on 91 original articles, employing productivity, impact, and collaboration indicators. SciVal facilitated data extraction, while VOSviewer 1.6.11 enabled the mapping of co-authorship networks and keyword co-occurrence. IBM SPSS Statistics 27 was used to determine correlations between variables of interest (Kendall’s rank correlation coefficient, statistically significant for p < 0.05).

**Results:** The 91 articles accumulated 2,197 citations (24.1/article), reflecting rising productivity. Most highly cited works (2019–2023) were published in Q1 journals. Dominant neurosurgical areas included neuro-oncology (25.4%) and education (20.9%), with AI applications focused on diagnostic accuracy (20.9%) and predictive tools (17.6%). Citations correlated with author numbers (p = 0.007). *World Neurosurgery* led in publications (Ndoc = 11), while *JAMA Network Open* had the highest citations/article (88.7). Author, institutional, and country productivity correlated strongly with citations (p < 0.001). Collaboration was universal (international: 29.7%, national: 53.8%, institutional: 16.5%).

**Conclusions:** The analyzed scientific output exhibited a marked quantitative growth trend and high citation rates, with a predominant focus on leveraging AI to enhance diagnostic accuracy, particularly in neuro-oncology. Publications were concentrated in specialized, high-impact journals and predominantly originated from authors and institutions in high-income, technologically advanced Northern Hemisphere countries, where scientific collaboration played a foundational role in driving research advancements.

## INTRODUCTION

The integration of artificial intelligence (AI) into medicine has catalyzed transformative advancements, ranging from diagnostic precision to personalized treatment strategies.^(1)^ Within neurosurgery, a discipline defined by its technical complexity and high-stakes decision-making, AI holds unprecedented potential to enhance surgical outcomes, optimize training paradigms, and streamline intraoperative workflows.^(2)^

Over the past decade, the proliferation of machine learning algorithms, computer vision systems, and predictive analytics has spurred a surge in scholarly output exploring AI applications in neurological surgery. Prior studies have examined AI’s role in specialized neurosurgical subdomains, such as spinal cord injuries, glioma research, and intracranial aneurysm management.^(3–5)^ El-Hajj *et al*.^(2)^ conducted a citation-driven analysis of the 50 most cited articles, while Levy *et al*.^(6)^ highlights the most impactful articles pertaining to machine learning in the field of neurosurgery. Similarly, reviews focus on technical challenges, such as endoscopic applications.^(7)^

However, despite this growth, the field lacks—to the best of our knowledge—a comprehensive bibliometric analysis to map its evolution, identify research priorities, and evaluate the impact of collaborative frameworks in the last decade. On one hand, while existing bibliometric analyses highlight specific applications or subfields, none provides a holistic overview of AI’s integration across all neurosurgical domains. On the other hand, some determinants of scholarly impact remain underexplored in this interdisciplinary field. Furthermore, rapid technological evolution necessitates an updated analysis of trends.

A systematic, decade-long assessment of AI-neurosurgery research is thus critical to contextualize advancements, address disparities in global contributions, and guide resource allocation for future innovation.

The objective of this study was to conduct a bibliometric analysis of Scopus-indexed (2014-2023) original articles at the intersection of AI and neurosurgery.

## METHOD

### Type of Study and Search Strategy

We conducted a bibliometric study to explore the topic. To achieve this, we applied the search strategy *(Neurosurg* OR “Neurologic* Surgery”) AND (“Artificial Intelligence” OR “AI” OR “Computer Reasoning” OR “Machine Intelligence” OR “Computational Intelligence” OR “Computer Vision System*” OR “Computer Knowledge Acquisition” OR “Computer Knowledge Representation*”)* in Scopus on May 25, 2024.

The exclusive use of Scopus is justified due to its comprehensive coverage and indexing of high-impact, peer-reviewed journals, ensuring the inclusion of relevant publications. Additionally, it offers advanced analytical tools, such as SciVal, which provide sophisticated metrics and visualizations essential for detailed research. Thus, Scopus serves as an authoritative, robust platform ideally suited for this study.

### Article Selection Process

From an initial pool of 764 results, we filtered by document type (original articles) and narrowed down the selection to 362. Following a thorough screening of titles and abstracts, the number of articles was further reduced to 276. Of these, we successfully retrieved 259 in full text, which were subsequently assessed for eligibility based on their publication year (2014-2023) and relevance to the proposed objective. Studies were retained if they explicitly addressed AI applications to neurosurgical practice, including but not limited to diagnostic accuracy, surgical planning, intraoperative decision-making, or training. Works addressing tangential topics were excluded. Ultimately, 91 articles were selected for inclusion in the study (**Figure 1**).

**Figure 1.**
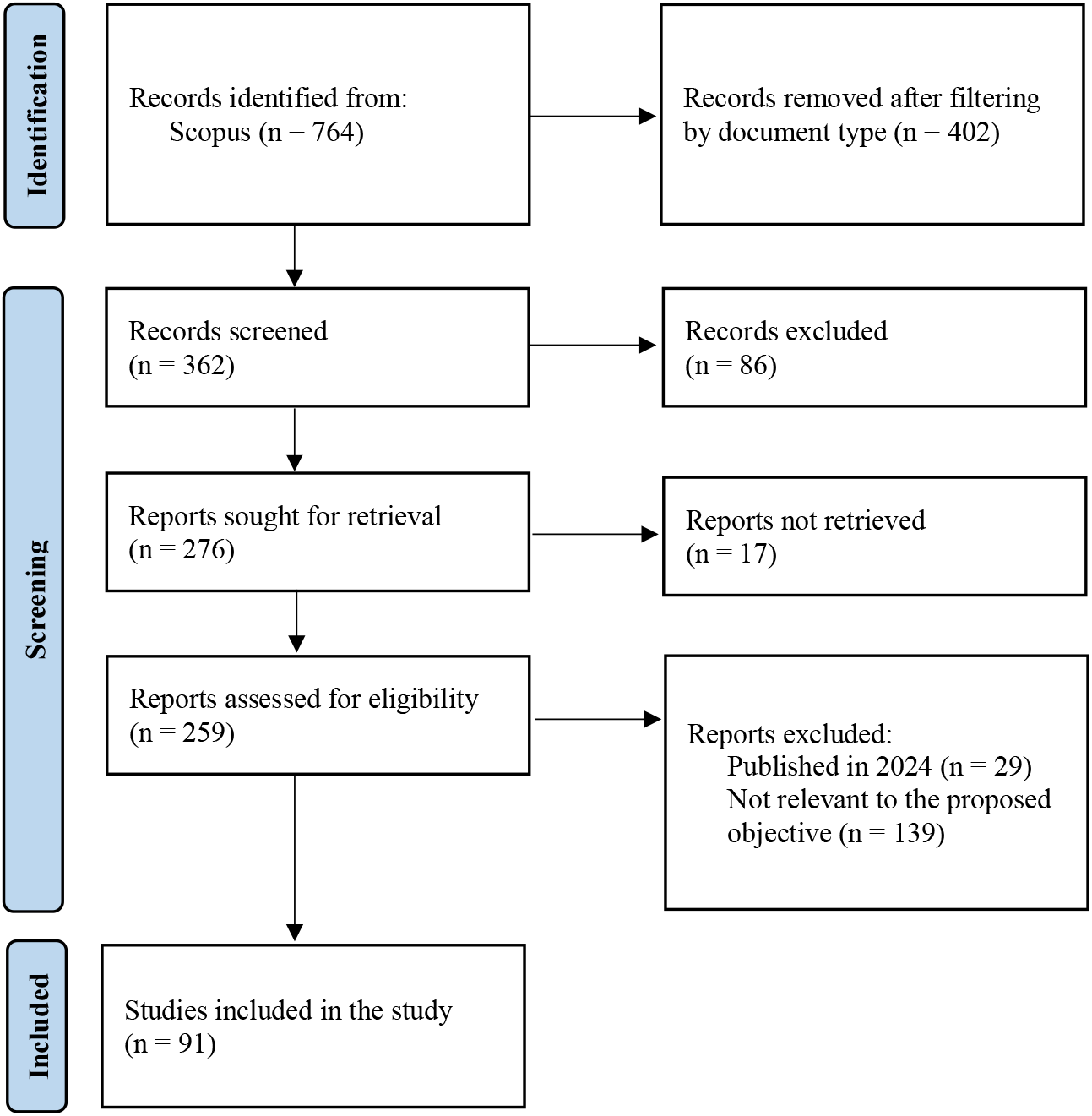
Flowchart of the search and study selection process. Source: modified version from Page et al.(8)

Original research articles were exclusively selected to ensure the analysis was grounded in empirical evidence and methodological rigor, as these studies provide primary data, validated results, and direct insights into AI applications within neurological surgery. By excluding other types of documents, such as reviews, editorials, and commentaries, we minimized bias from secondary interpretations or speculative discussions, focusing instead on novel contributions that advance the field.

### Data Extraction, Metric Indicators and Network Mapping

The selected articles were added to a library in Zotero 6.0.36 and subsequently exported in comma-separated values (CSV) format. This file was then uploaded to the bibliometric suite SciVal for data extraction on November 28, 2024. The metric indicators studied are presented in **Figure 2**. VOSviewer 1.6.11 was utilized to map co-authorship networks and keyword co-occurrence, employing fractional counting with a threshold of three and weighting based on occurrences. The website https://www.mapchart.net/ was utilized to generate a world map illustrating the distribution of the most productive countries.

**Figure 2.**
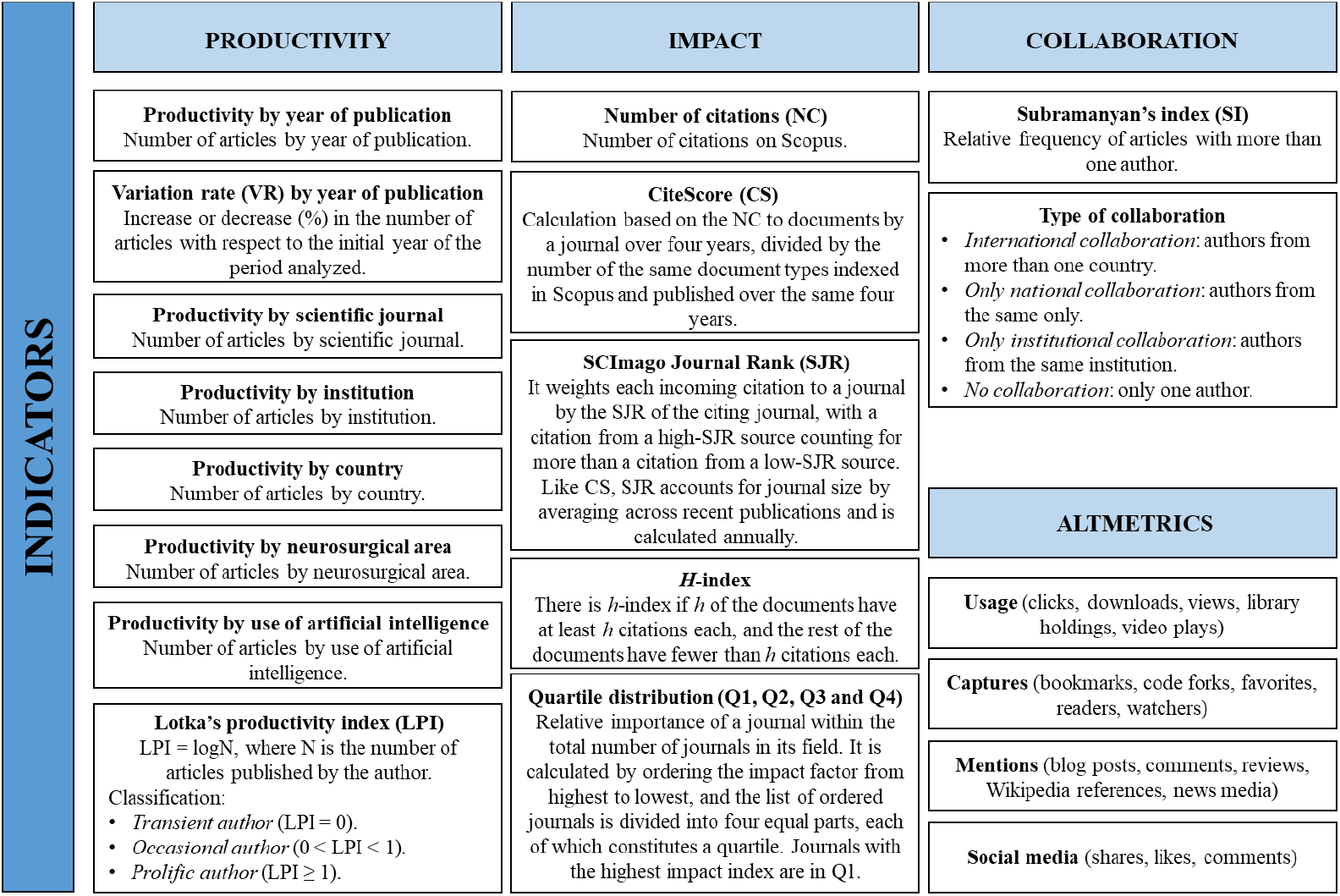
Metric indicators. Source: elaborated by the authors.

### Statistical Analysis

The statistical analysis was conducted using IBM SPSS Statistics 27. The Kendall’s Tau correlation test was employed to identify correlations between variables after confirming the non-normal distribution via the Kolmogorov-Smirnov-Lilliefors test. A p-value of less than 0.05 was considered statistically significant.

## RESULTS

### Productivity and impact

#### Article-level metrics

The 91 original articles garnered 2,197 citations, averaging 24.1 citations per article. As illustrated in **Figure 3**, both the number of documents (Ndoc) and the NC have exhibited an upward quantitative trend. No scientific output was recorded in the years 2014 and 2015. The average NC per year was 219.7.

**Figure 3.**
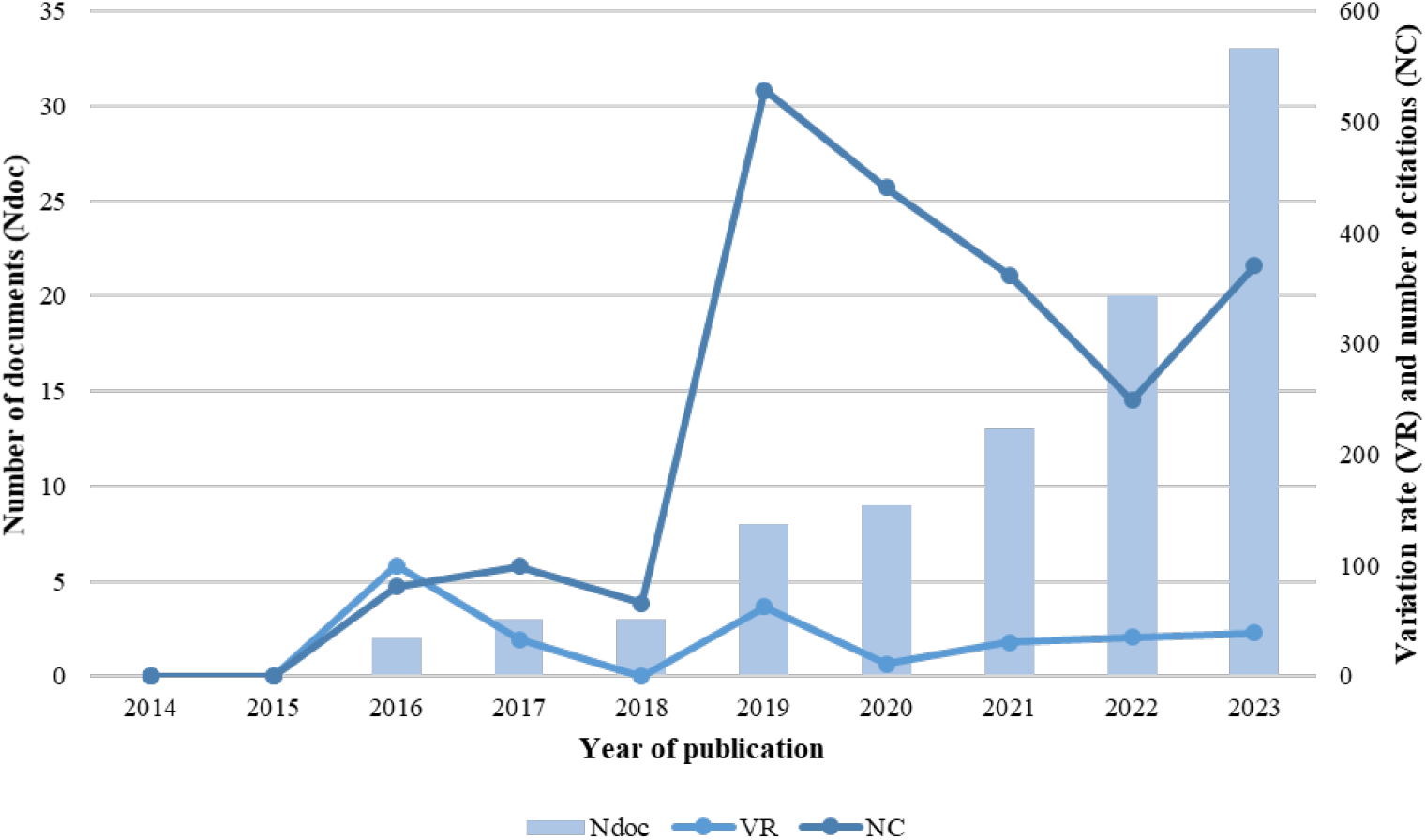
Productivity, VR and NC by year of publication.

Eighty percent of the ten most cited articles were published between 2019 and 2023, with 2019 producing the highest number of these publications. Their NC ranged from 60 to 150, while the CS shown by their journal in the year they were published varied from 1.7 to 15.1. Ninety percent of these articles are in first-quartile journals. Alternative metric data, particularly captures and citation indexes, have been recorded for all of them (**Table 1**).

**Table 1.** Top 10 most cited articles.

| Rank | Year | Title | NC | CS | First author | Journal (Quartile\|SJR 2023) | Altmetrics (PlumX metrics*) |
| --- | --- | --- | --- | --- | --- | --- | --- |
| 1 | 2020 | The virtual operative assistant: An explainable artificial intelligence tool for simulation-based training in surgery and medicine | 150 | 5.3 | Mirchi N | *PLoS ONE* (Q1\|0.84) | Captures: 424<br>News<br>Mentions: 1<br>Citations<br>Indexes: 1<br>Policy<br>Citations: 1<br>Shares, Likes and<br>Comments: 6 |
| 2 | 2023 | Performance of ChatGPT, GPT-4, and Google Bard on a Neurosurgery Oral Boards Preparation Question Bank | 107 | 8.2 | Ali R | *Neurosurgery* (Q1\|1.31) | Captures: 192<br>Citations<br>Indexes: 59 |
| 3 | 2019 | Development of Machine Learning Algorithms for Prediction of 30-Day Mortality after Surgery for Spinal Metastasis | 104 | 6.2 | Karhade AV | *Clinical Neurosurgery* (N/A) | Captures: 146<br>Citation Indexes: 90 |
| 4 | 2019 | Machine Learning Identification of Surgical and Operative Factors Associated with Surgical Expertise in Virtual Reality Simulation | 101 | 1.7 | Winkler-Schwartz A | *JAMA Network Open* (Q1\|3.41) | Captures: 252<br>News Mentions: 7<br>References: 1<br>Citations Indexes: 1<br>Shares, Likes and Comments: 82 |
| 5 | 2019 | Artificial Intelligence Based Hierarchical Clustering of Patient Types and Intervention Categories in Adult Spinal Deformity Surgery: Towards a New Classification Scheme that Predicts Quality and Value | 98 | 5.1 | Ames CP | *Spine* (Q1\|1.22) | Captures: 198<br>News Mentions: 1<br>Citations Indexes: 67 |
| 6 | 2019 | Artificial Intelligence Distinguishes Surgical Training Levels in a Virtual Reality Spinal Task | 82 | 6.9 | Bissonnette V | *Journal of Bone and Joint Surgery - American Volume* (Q1\|1.71) | Captures: 192<br>Citations Indexes: 59 |
| 7 | 2021 | An artificial intelligence framework for automatic segmentation and volumetry of vestibular schwannomas from contrast-enhanced T1-weighted and high-resolution T2-weighted MRI | 70 | 8 | Shapey J | *Journal of Neurosurgery* (Q1\|1.17) | Captures: 94<br>Citations Indexes: 72 |
| 8 | 2017 | A machine learning approach for real-time modelling of tissue deformation in image-guided neurosurgery | 65 | 4.9 | Tonutti M | *Artificial Intelligence in Medicine* (Q1\|1.72) | Captures: 219<br>Patent Family Citations: 2<br>Citations Indexes: 12<br>Shares, Likes and Comments: 2 |
| 9 | 2022 | Effect of artificial intelligence tutoring vs expert instruction on learning simulated surgical skills among medical students a randomized clinical trial | 65 | 15.1 | Fazlollahi AM | *JAMA Network Open* (Q1\|3.41) | Captures: 419<br>Citations: 1<br>Citations Indexes: 1<br>Policy<br>Citations: 1<br>Shares, Likes and<br>Comments: 6 |
| 10 | 2018 | Automatic evaluation of traumatic brain injury based on terahertz imaging with machine learning | 60 | 6.8 | Shi J | *Optics Express* (Q1\|1) | Captures: 81<br>Citations Indexes: 59 |

\*Extracted from Scopus.
NC: number of citations. CS: CiteScore. SJR: SCImago Journal Rank. N/A: not assigned.

The articles were categorized into the following neurosurgical areas: endoscopic (2.2%), trauma (4.4%), functional (7.7%), vascular (8.8%), neuro-oncology (15.4%), spine (16.5%), education (20.9%), and miscellaneous (24.2%). The most frequent uses of AI observed in neurosurgery included the enhancement of diagnostic accuracy (20.9%), its application as a prediction tool (17.6%), and the analysis of neurosurgical performance, skills, and expertise (9.9%), as shown in **Figure 4**.

**Figure 4.**
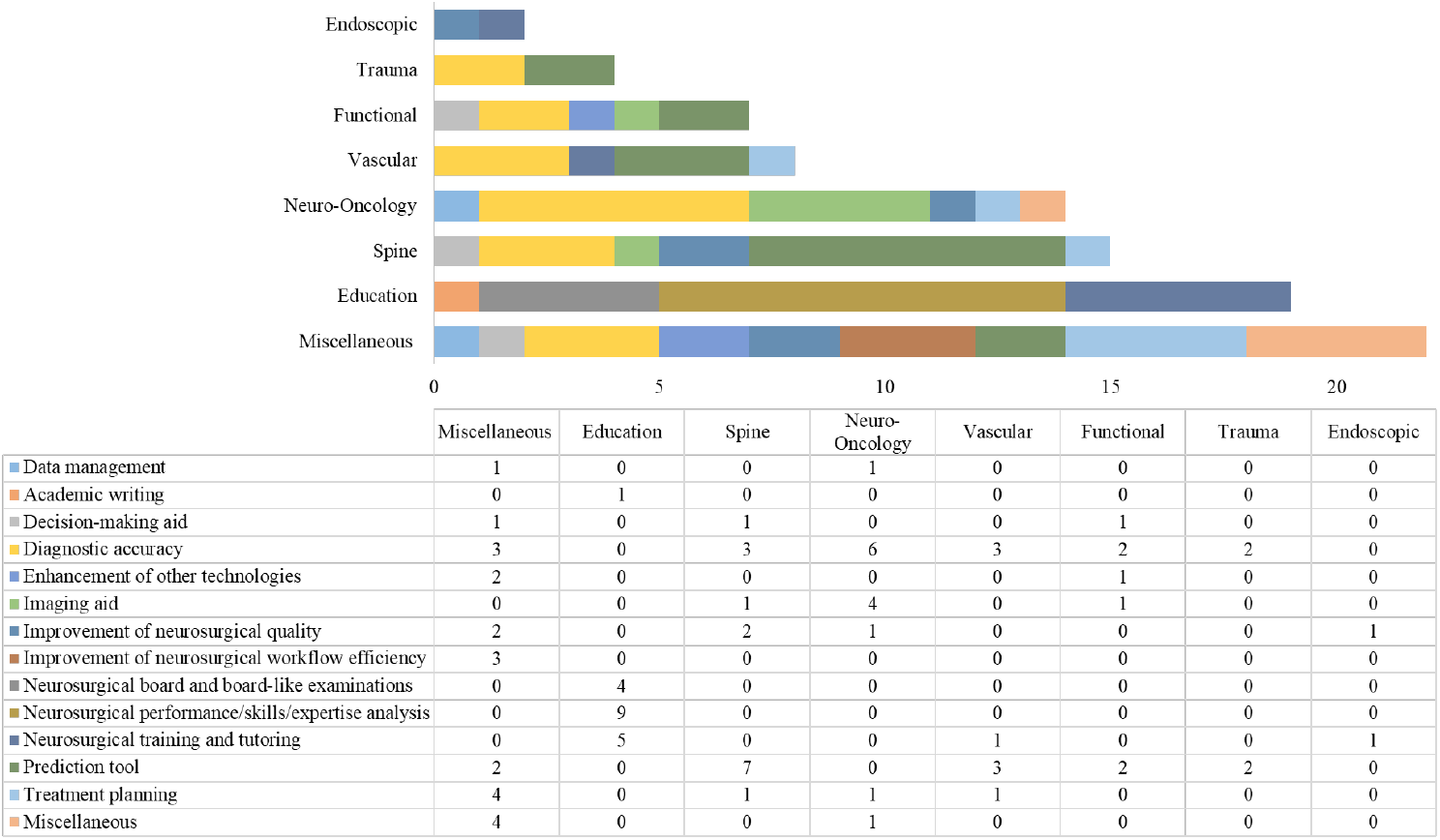
Productivity by surgical area and use of AI.

Based on the keyword co-occurrence map analysis, we identified 176 items, which were grouped into seven distinct clusters. The map revealed a substantial network of interconnections, with 6,150 links established between the items. The cumulative link strength of these connections was quantified at 776.5, indicating a robust level of inter-item association. This visualization underscores the relationships and trends in research topics, helping to identify key areas of focus and potential collaborations in these fields (**Figure 5**).

**Figure 5.**
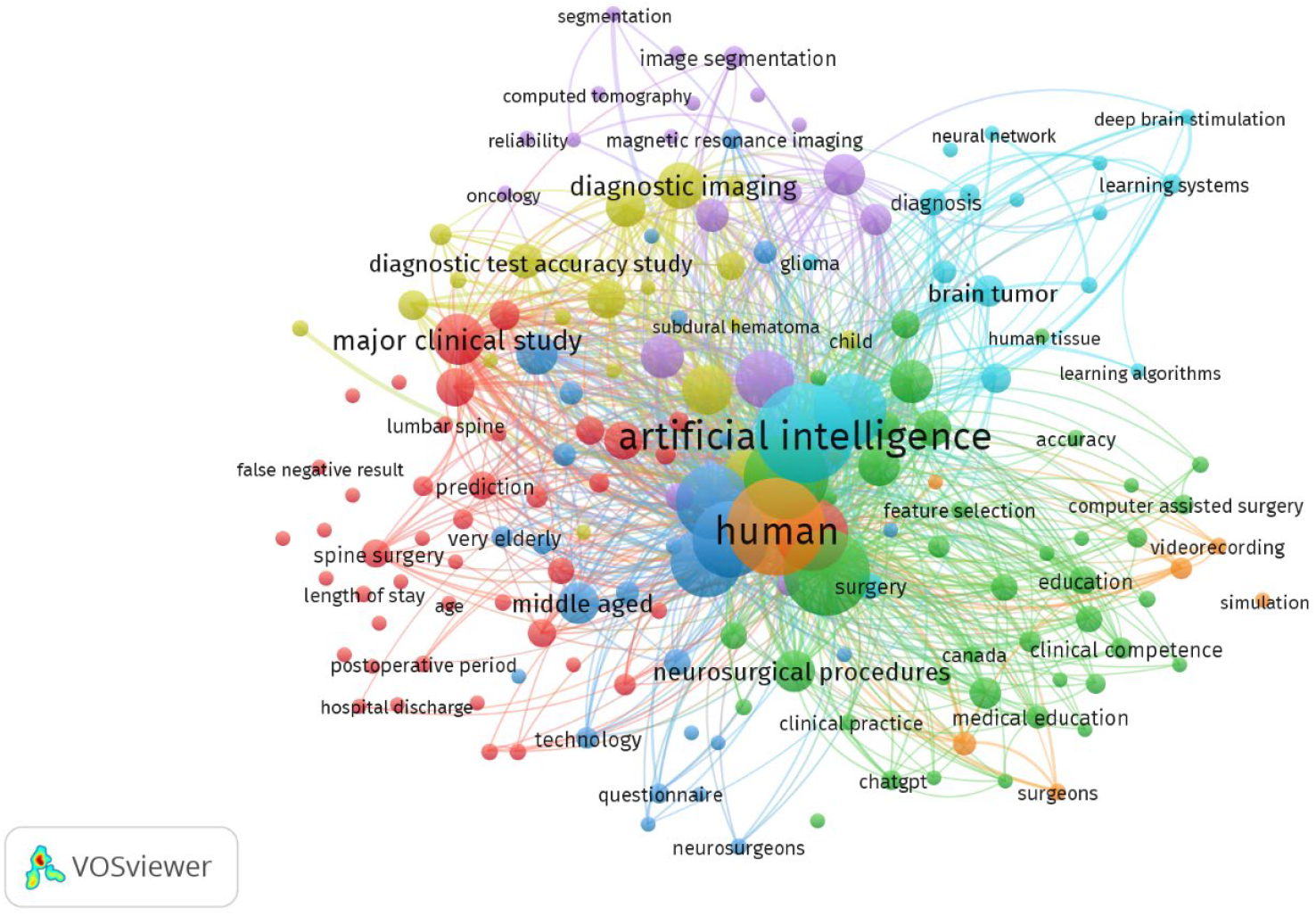
Map of keyword co-occurrence.

**Figure 6** shows the top 10 topics with the highest Ndoc.

**Figure 6.**
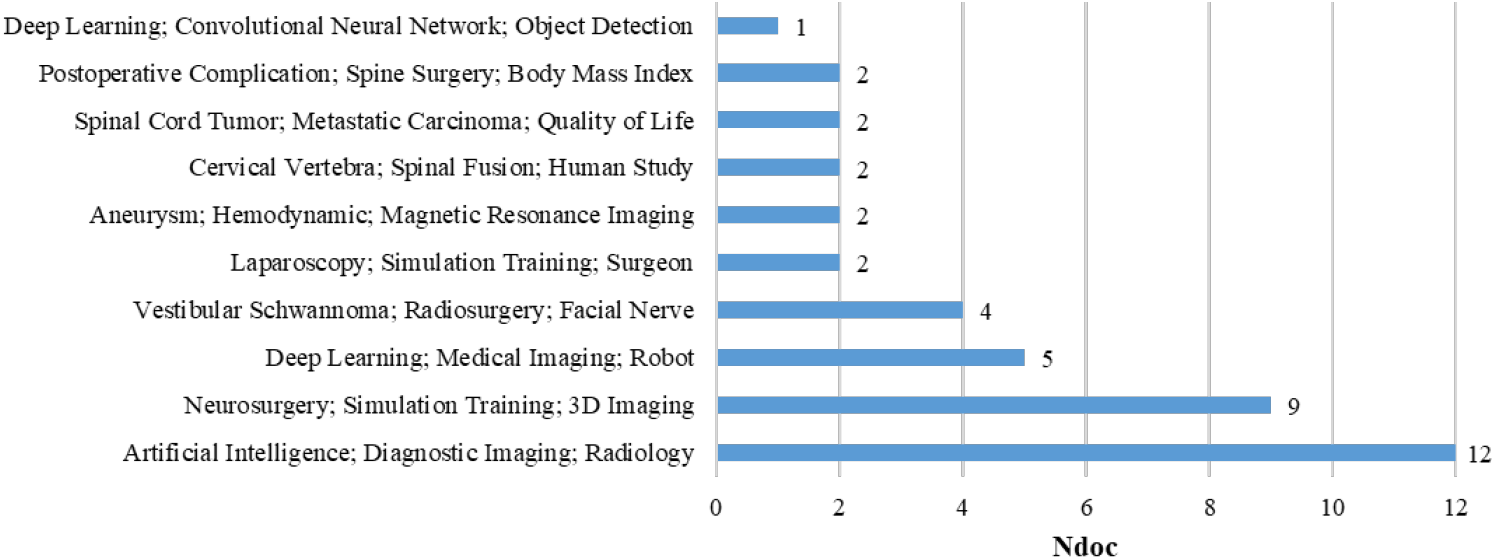
Top 10 topics with the highest Ndoc. Ndoc: number of documents.

The analysis revealed a statistically significant positive correlation between the NC and the number of authors (p = 0.007), indicating that papers with more authors tend to receive more citations. However, no significant correlations were found between the number of citations and the number of institutions (p = 0.102) or the number of countries (p = 0.235) involved in the research.

#### Journal-level metrics

The analysis of top journals reveals that *World Neurosurgery* leads in the number of documents with 11 publications, but has a moderate NC per publication of 27.2. In contrast, *JAMA Network Open, Neurosurgery* and *PLoS ONE*, though having only three publications each, stand out with the highest NC per publication, reflecting strong citation influence. Most of the top journals are in the first quartile and are specifically dedicated to neurosurgery, underscoring their high quality and impact in their respective fields. Collectively, the top 10 most productive journals account for 47% of all the articles studied and 57% of all citations (**Table 2**).

**Table 2.** Top 10 journals with the highest number of publications.

| Rank | Journal | Ndoc | NC | NC per publication | Quartile | SJR 2023 | Field-specific |
| --- | --- | --- | --- | --- | --- | --- | --- |
| 1 | *World Neurosurgery* | 11 | 299 | 27.2 | Q2 | 0.65 | Yes |
| 2 | *Neurosurgical Focus* | 6 | 26 | 4.3 | Q1 | 1.31 | Yes |
| 3 | *Frontiers in Surgery* | 4 | 49 | 12.3 | Q2 | 0.44 | No |
| 4 | *Journal of Neurosurgery* | 4 | 134 | 33.5 | Q1 | 1.17 | Yes |
| 5 | *Neurosurgery* | 3 | 171 | 57 | Q1 | 1.31 | Yes |
| 6 | *JAMA Network Open* | 3 | 266 | 88.7 | Q1 | 3.41 | No |
| 7 | *Operative Neurosurgery* | 3 | 16 | 5.3 | Q2 | 0.58 | Yes |
| 8 | *PLoS ONE* | 3 | 165 | 55 | Q1 | 0.84 | No |
| 9 | *Scientific Reports* | 3 | 34 | 11.3 | Q1 | 0.9 | No |
| 10 | *Spine Journal* | 3 | 93 | 31 | Q1 | 1.8 | Yes |

Ndoc: number of documents. NC: number of citations. SJR: SCImago Journal Rank.

A statistically significant positive correlation was found between the Ndoc in a journal and the NC (p < 0.001). This indicates that journals publishing more documents tend to receive higher citation counts.

#### Author-level metrics

Among the most productive authors, predominantly from Canada and particularly McGill University, Nicole Ledwos and Nykan Mirchi lead with high citation counts per publication (NC = 438), and Joseph Hasbrouck Schwab stands out with the highest h-index (h = 49), indicating substantial impact in his field. Institutions in the United States, including Massachusetts General Hospital and the University of Southern California, also feature prominently. The ocassional nature of all top authors, as per the LPI classification, is noteworthy (**Table 3**).

**Table 3.** Top 10 most productive authors.

| Rank | Author's name | Affiliation | Country | Ndoc | h-index | NC | NC per publication | Type of author (LPI) |
| --- | --- | --- | --- | --- | --- | --- | --- | --- |
| 1 | Ledwos, Nicole | Centre for Addiction and Mental Health | Canada | 7 | 13 | 438 | 62.6 | Occasional |
| 2 | Mirchi, Nykan | McGill University | Canada | 7 | 12 | 438 | 62.6 | Occasional |
| 3 | Winkler-Schwartz, Alexander | McGill University | Canada | 7 | 18 | 419 | 59.9 | Occasional |
| 4 | Yilmaz, Recai | McGill University | Canada | 7 | 9 | 362 | 51.7 | Occasional |
| 5 | Del Maestro, Rolando F. | McGill University | Canada | 6 | 43 | 429 | 71.5 | Occasional |
| 6 | Marcus, Hani Joseph | University College London | United Kingdom | 5 | 34 | 146 | 29.2 | Occasional |
| 7 | Bissonnette, Vincent | McGill University | Canada | 4 | 9 | 343 | 85.8 | Occasional |
| 8 | Karhade, Aditya Vishwas | Massachusetts General Hospital | United States | 4 | 33 | 239 | 59.8 | Occasional |
| 9 | Pangal, Dhiraj J. | University of Southern California | United States | 4 | 12 | 58 | 14.5 | Occasional |
| 10 | Schwab, Joseph Hasbrouck | Massachusetts General Hospital | United States | 4 | 49 | 239 | 59.8 | Occasional |

Ndoc: number of documents. NC: number of citations. LPI: Lotka's productivity index.

Authors with more publications tend to receive more citations, as evidenced by the statistically significant positive correlation found between these variables (p < 0.001).

#### Institution-level metrics

**Table 4** reveals that University College London and its affiliated NHS Foundation Trust are leading contributors, each producing nine publications with a notable citation count per publication. Institutions such as Massachusetts General Hospital and McGill University have fewer publications but exceptionally high citation counts per publication, indicating a significant impact in their fields. The analysis also highlights the diversity in geographical representation, with top-performing institutions spanning the United Kingdom, Canada, the United States, and China.

**Table 4.**
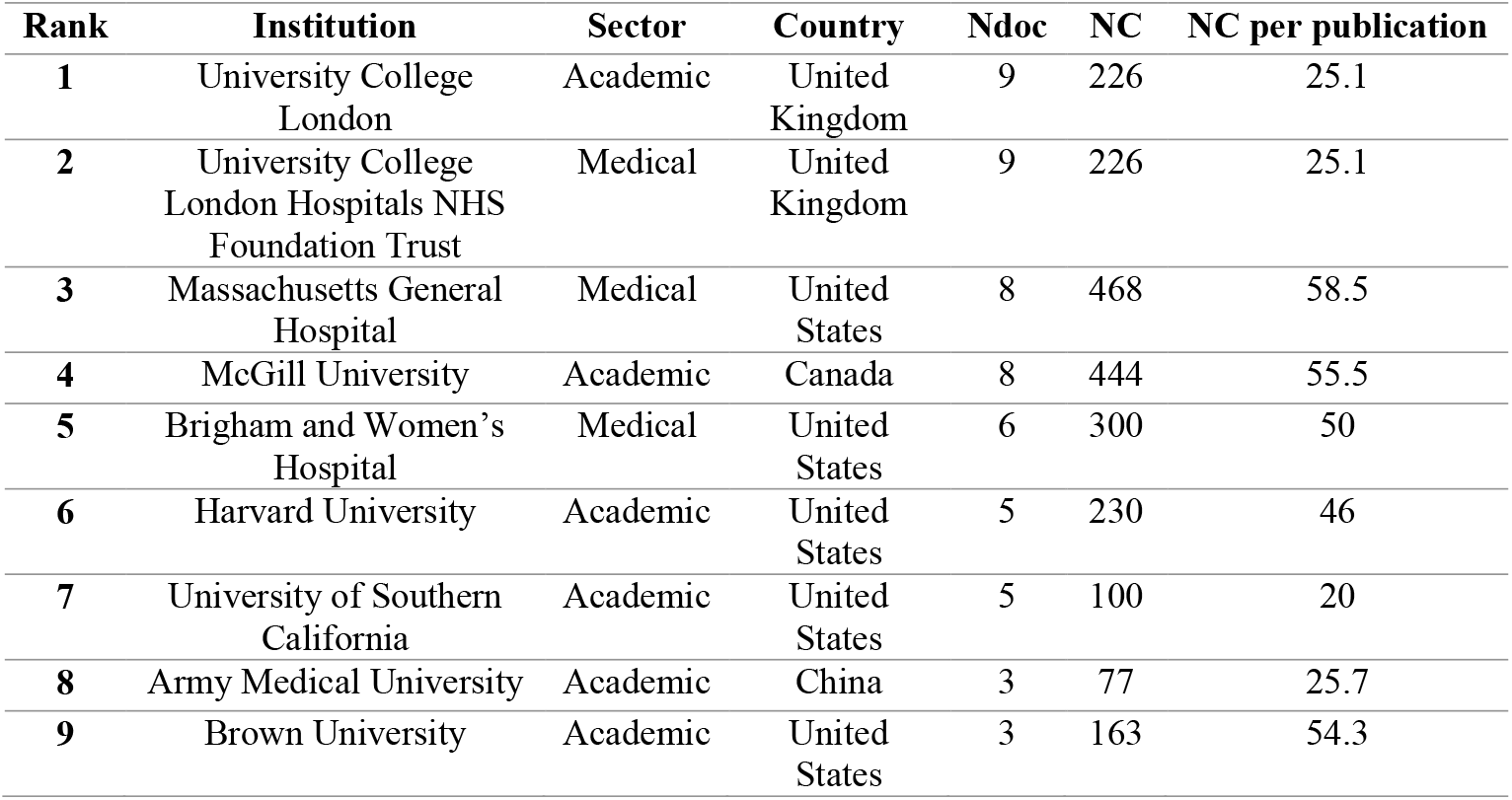
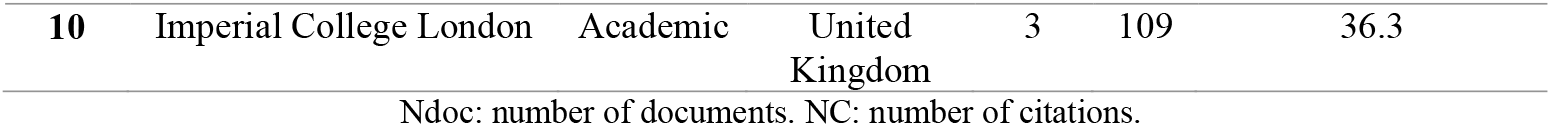
Top 10 most productive institutions.

| Rank | Institution | Sector | Country | Ndoc | NC | NC per publication |
| --- | --- | --- | --- | --- | --- | --- |
| 1 | University College London | Academic | United Kingdom | 9 | 226 | 25.1 |
| 2 | University College London Hospitals NHS Foundation Trust | Medical | United Kingdom | 9 | 226 | 25.1 |
| 3 | Massachusetts General Hospital | Medical | United States | 8 | 468 | 58.5 |
| 4 | McGill University | Academic | Canada | 8 | 444 | 55.5 |
| 5 | Brigham and Women's Hospital | Medical | United States | 6 | 300 | 50 |
| 6 | Harvard University | Academic | United States | 5 | 230 | 46 |
| 7 | University of Southern California | Academic | United States | 5 | 100 | 20 |
| 8 | Army Medical University | Academic | China | 3 | 77 | 25.7 |
| 9 | Brown University | Academic | United States | 3 | 163 | 54.3 |
| 10 | Imperial College London | Academic | United Kingdom | 3 | 109 | 36.3 |

Ndoc: number of documents. NC: number of citations.

Institutions with higher publication output tend to attract more citations. This is supported by the statistically significant positive correlation observed between the Ndoc and the NC (p < 0.001).

#### Country-level metrics

Figure 7. highlights key nations such as the United States, the United Kingdom and China, indicating their significant contributions to global research. The visual representation reveals a concentration of highly productive countries in the Northern Hemisphere, particularly in North America and Europe, as well as in parts of Asia. This distribution underscores the prominence of these regions in academic research on neurosurgery and AI.

A statistically significant positive correlation was found between the Ndoc produced by a country and the citation count (p < 0.001). This suggests that countries with a higher volume of publications tend to receive more citations.

**Figure 7.**
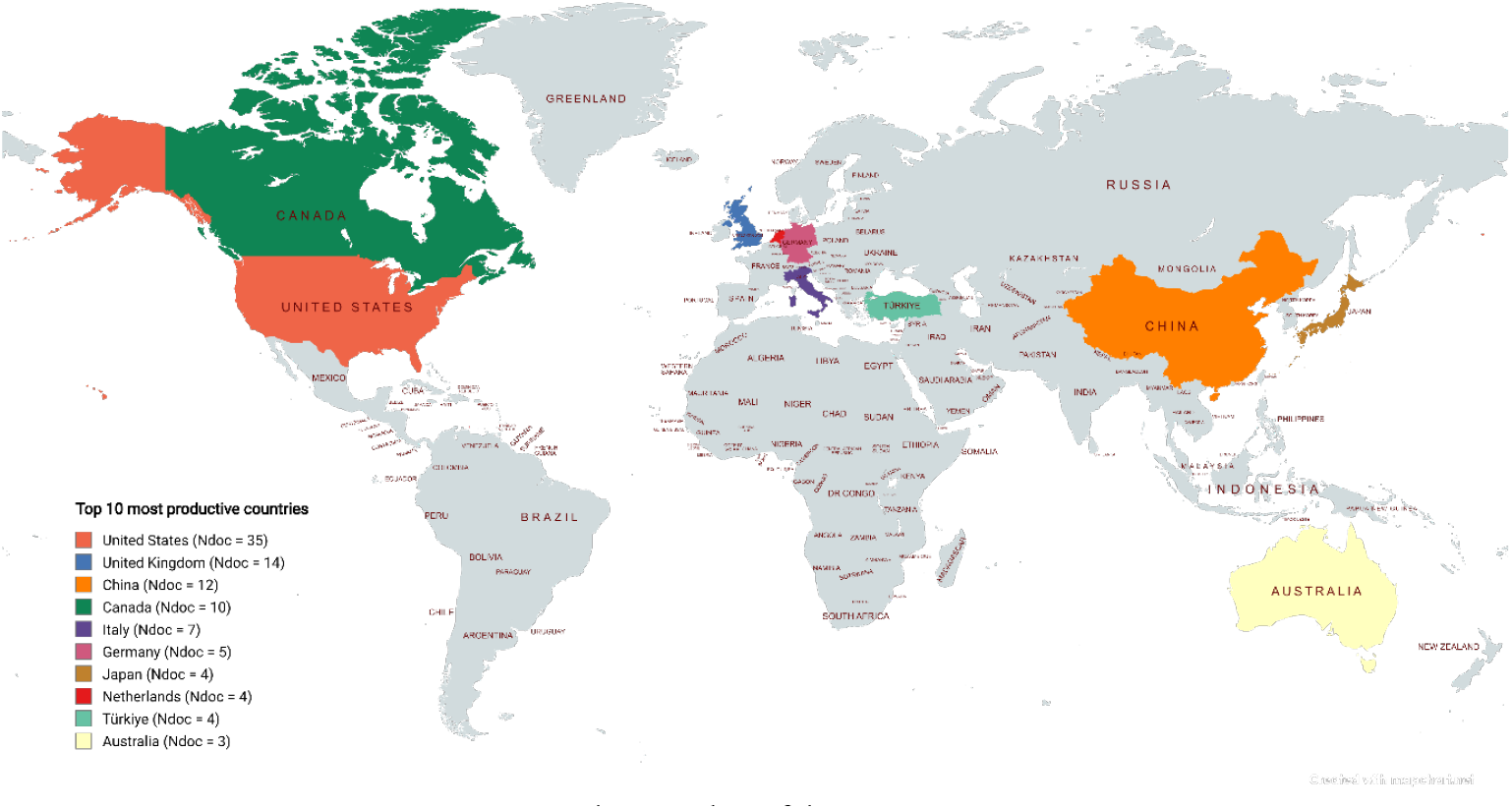
Top 10 most productive countries. Ndoc: number of documents.

### Collaboration

The analysis of scholarly output and collaboration types reveals that international collaborations, despite accounting for only 29.7% of the total scholarly output, achieve a high impact with an average of 29.4 citations per publication. National collaborations dominate in quantity, yet they yield a lower average of citations per publication. Institutional collaborations, while less common, have a similar citation impact to international ones. Notably, single authorship shows no output or citations (SI = 100%), underscoring the importance of collaborative efforts in enhancing the visibility and impact of academic research.

Figure 8. presents the largest co-authorship network identified (links: 15; total link strength: 19). It is notable that this network pertains to Canadian authors, who top the list of the most productive authors, as previously shown in **Table 5**.

## DISCUSSION

Over the past decade, the intersection of AI and neurosurgery has emerged as a catalytic axis for medical innovation, as evidenced by a marked increase in scientific output and the discernible scholarly influence of studies within this domain.

**Table 5.** Types of collaboration.

| Type of collaboration | Ndoc | Ndoc % | NC | NC per publication |
| --- | --- | --- | --- | --- |
| International collaboration | 27 | 29.7% | 794 | 29.4 |
| Only national collaboration | 49 | 53.8% | 979 | 20 |
| Only institutional collaboration | 15 | 16.5% | 424 | 28.3 |
| No collaboration | 0 | 0% | 0 | 0 |

Ndoc: number of documents. NC: number of citations.

**Figure 8.**
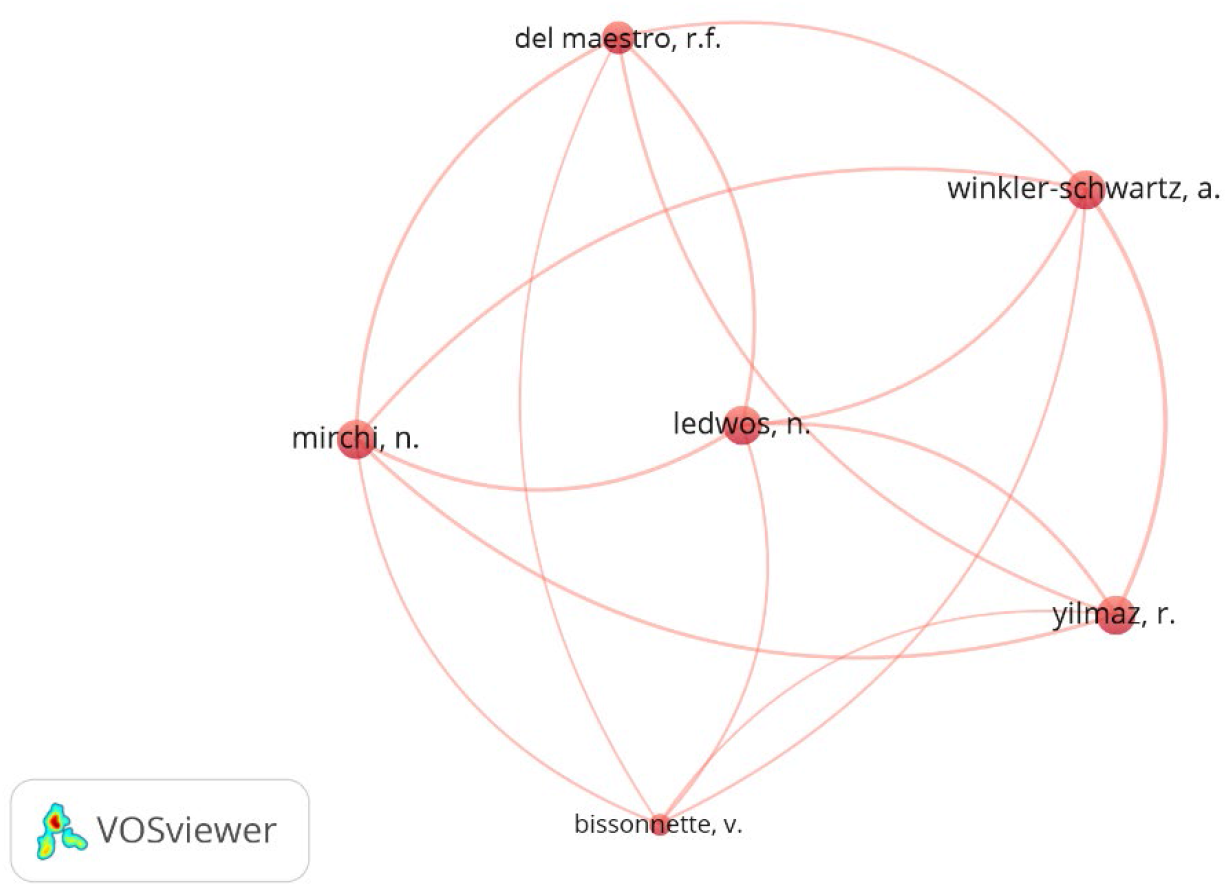
Largest co-authorship network.

### Temporal Trends in Scientific Productivity

The analytical findings delineate a pronounced trajectory in scientific productivity, characterized by a complete absence of publications in 2014 and 2015, followed by a progressive surge commencing in 2016. Notably, 80% of the ten most frequently cited works within the dataset were disseminated between 2019 and 2023. This temporal pattern corresponds to a prototypical three-year maturation cycle, wherein methodological refinement of computational models precedes their subsequent translation into clinical validation—a phenomenon extensively corroborated in prior literature.^(2–4,9–13)^ These trends collectively underscore the exponential integration of AI into medical and, particularly, neurosurgical research, a trajectory further paralleled by escalating scholarly engagement. The intensifying academic interest is quantitatively evidenced by robust citation metrics, with the analyzed corpus accruing 2,197 citations in the present study and cumulative citations exceeding 2,000 when contextualized alongside antecedent investigations.^(5,11,13,14)^

The year 2019 is identified as the pinnacle of high-impact scholarly contributions within the analyzed corpus, contrasting with prior studies that designate 2021^(2,4)^ and 2022^(3,5)^ as periods of maximal publication volume and citation frequency. This discrepancy likely stems from methodological disparities in article selection criteria. For instance, the present analysis prioritized original research articles indexed in Scopus (2014–2023), whereas El-Hajj *et al*.^(2)^ employed a citation-driven approach focused on the 50 most cited works in Web of Science without temporal constraints. Furthermore, comparative investigations(3–5) centered on specialized neurosurgical subdomains—spinal cord neural injury, glioma research, and intracranial aneurysm management—thereby contextualizing observed variances in productivity trends. A consistent observation across studies is the decline in both publication output and citation metrics for recent years (2023–2024),^(3,5,11,15)^ a phenomenon potentially attributable to temporal proximity to publication deadlines and indexing lags.^(11)^

### Thematic Focus and AI-Driven Innovations

Our analysis identifies eight neurosurgical domains integrating AI, with predominant applications in education (20.9%), spinal surgery (16.5%), and neuro-oncology (15.4%). This distribution aligns closely with the findings of El-Hajj *et al*.,^(2)^ wherein spinal surgery and neuro-oncology similarly dominate (26% and 18%, respectively, among the 50 most cited works), alongside endovascular neurosurgery (24%). However, neurosurgical education emerges as a prominent focus in our dataset but not in the comparative bibliometric analysis (20.9% vs. 4%), a discrepancy likely attributable to the nascent stage of AI-driven educational methodologies in neurosurgery, which have not yet attained critical scholarly traction or clinical integration. Within this educational subfield, novel opportunities are emerging for developing standardized training protocols and immersive simulation systems, which may mitigate procedural error rates and enhance surgical learning curves in the near term.^(16)^

Endoscopic surgery, despite its significance, exhibits minimal representation in both the present analysis and the study by El-Hajj *et al*.,^(2)^ accounting for 2.2% and 2% of cases, respectively. This underrepresentation may be attributed, as posited by prior investigations,^(1)^ to persistent technical barriers in achieving high-fidelity algorithmic reliability for endoscopic applications, alongside the inherent complexity and resource-intensive demands of curating and annotating endoscopic surgical datasets^(7)^—factors constraining the development of robust training databases.

Regarding AI applications, both our findings and those of El-Hajj *et al*.^(2)^ converge in identifying diagnostic accuracy enhancement (20.9% vs. 28% of studies, respectively) and predictive analytics (17.6% vs. 32%) as predominant use cases. This parity underscores their enduring scientific priority within AI-driven neurosurgical innovation.

### Stratified Analysis of Scholarly Output and Its Impact

At the journal level, productivity analysis reveals a pronounced predominance of publications in high-impact factor journals, underscoring the field’s prioritization within the scientific community. Leading venues include *JAMA Network Open* (88.7 citations per article), *Neurosurgery* (57 citations per article), and *PLoS ONE* (55 citations per article), with *World Neurosurgery* demonstrating moderate citation metrics (33.5 citations per article) despite higher publication volume. These findings align with prior observations by El-Hajj *et al*.,^(2)^ wherein *JAMA Network* similarly exhibited the highest impact factor. Notably, journals with elevated document counts (e.g., *World Neurosurgery*) exhibit disproportionately lower citation indices per article compared to high-impact, low-volume counterparts. This dichotomy reflects the inherent trade-off between scholarly output volume and methodological rigor, suggesting that pioneering AI-neurosurgical research remains constrained by the resource-intensive nature of high-impact study design.^(17,18)^

Author-level productivity analysis reveals a pronounced concentration of researchers affiliated with elite academic institutions, notably McGill University and other leading Canadian centers. This finding corroborates the work of Ponce *et al*.,^(19)^ whose bibliometric study identified Canadian neurosurgery departments—particularly those at the University of Toronto and McGill University—as dominant contributors to high-impact scholarly output within the discipline. When evaluated through LPI, none of the analyzed authors exceeded 10 publications, with even the most prolific individuals categorized as occasional contributors. This trend likely reflects the nascent stage of AI-neurosurgical research, wherein the field’s rapid evolution and interdisciplinary demands impede the emergence of sustained scholarly productivity. Leading contributors include Nicole Ledwos and Nykan Mirchi, each with 7 publications and a cumulative citation count of 438, yielding an average of 62.6 citations per article. Notably, Alexander Winkler-Schwartz, despite a lower NC, has authored two articles ranked among the top 50 most cited works in this domain.^(2)^

Institutional-level analysis reveals a marked divergence between publication volume and scholarly impact. Institutions such as University College London and its affiliated NHS Foundation Trust, for instance, rank among the top six centers in the application of large language models to medicine (Ndoc=15),^(14)^ with nine publications demonstrating high publication output. However, their citation metrics remain comparatively modest at 25.1 citations per document. In contrast, institutions like Massachusetts General Hospital and McGill University—each with eight publications—exhibit disproportionately higher citation indices (58.5 and 55.5 citations per document, respectively). Notably, McGill University contributes three articles to the 50 most cited works in neurosurgery and AI.^(2)^ This disparity likely reflects systemic inequities in resource allocation, including the heightened allocation of temporal and financial investments, the strategic cultivation of inter-institutional collaborative networks,^(20,21)^ and the preexisting academic prominence of affiliated researchers—factors that critically determine institutional scientific influence independent of productivity metrics.

Geospatial analysis of scholarly output demonstrates a pronounced concentration in the United States (Ndoc = 35), the United Kingdom (Ndoc = 14), and China (Ndoc = 12), mirroring established trends in both global neurosurgical research(22) and AI applications in Medicine.^(14)^ Notably, the United States exerts disproportionate influence, contributing 22 of the 50 most cited articles on AI in neurosurgery, followed by Canada with 5 articles^(2)^—a nation ranked fourth in quantitative output (Ndoc = 10) within our dataset. This discrepancy between publication volume and scholarly impact is particularly evident when comparing Canada’s high-impact contributions to nations with greater quantitative output but lower citation metrics. The observed Canadian prominence may be linked to the methodological rigor and innovation prioritization of its neurosurgical training programs,^(23,24)^ which appear to favor quality over quantity in research output.

The disproportionately high scientific output observed in these nations arises from four interdependent determinants: substantial public and private investment in research and development, a critical mass of specialized human capital within neurosurgical and computational disciplines, institutional prioritization of high-risk, high-reward innovation pipelines, and regulatory frameworks incentivizing translational research through targeted funding mechanisms.^(18,24–26)^ The synergistic interplay of these factors—specialized expertise, infrastructural resourcing, and policy-driven financial support—cultivates a self-reinforcing cycle of innovation, positioning these ecosystems as global epicenters for AI-driven neurosurgical advancements.

### Collaboration Dynamics and Knowledge Networks

This analysis underscores the critical role of international collaborative frameworks in circumventing logistical and regulatory constraints, fostering complementary expertise and infrastructural synergies, and securing extramural funding streams.^(27–30)^

Evaluation of co-authorship networks reveals a notable trend: publications stemming from international (29.4 citations/publication) and institutional (28.3 citations/publication) collaborations demonstrate statistically comparable citation yields, both substantially exceeding those of national collaborations (20 citations/publication). Contrary to conventional expectations, this parity suggests that within AI-neurosurgical research, strategic alliances among elite national institutions—through the aggregation of cutting-edge computational resources, multidisciplinary teams (integrating neurosurgical, engineering, and data science expertise), and institutional prestige—can approximate the scholarly impact typically associated with global consortia. Such configurations effectively attenuate the anticipated disparity between local and international collaboration scales, redefining the relationship between geographic scope and academic influence in this interdisciplinary domain.

### Limitations

Reliance on Scopus as the sole database may have excluded relevant articles indexed in other repositories, potentially introducing selection bias. The search strategy, while comprehensive, might not have captured all AI-related terminology. Temporal limitations arise from the cutoff year (2023), as indexing delays may underrepresent recent high-impact works. The focus on original articles excluded reviews and commentaries, which might offer complementary perspectives. Analytical tools like VOSviewer and SciVal, though robust, have inherent algorithmic constraints in network mapping and citation interpretation. Geographic and institutional productivity trends may reflect Scopus’s coverage bias rather than true global research activity. Finally, quantitative metrics such as citation counts do not fully assess clinical relevance or innovation, and observed correlations may oversimplify multifaceted determinants of scholarly impact.

### Contributions

This study offers a comprehensive bibliometric analysis that delineates the evolving landscape of AI integration in neurosurgery over the past decade, providing critical insights into productivity trends, research priorities, and collaboration dynamics. By identifying dominant AI application areas, the work highlights emerging frontiers and guides future research investments. The analysis reveals the outsized scholarly impact of international and institutional collaborations compared to national efforts, underscoring the strategic value of cross-disciplinary and cross-border partnerships. By mapping leading journals, authors, institutions, and countries, the study aids in identifying knowledge networks and fostering global research synergies. Furthermore, the correlations observed reinforce the importance of collaborative frameworks in driving innovation.

These contributions collectively enhance understanding of AI’s transformative role in neurosurgery, offering actionable insights for policymakers, educators, and researchers to optimize resource allocation, prioritize translational studies, and address under-explored domains.

### Final considerations

This bibliometric analysis underscores the transformative role of AI in reshaping neurosurgical research and practice over the past decade. The analyzed scientific output exhibited a marked quantitative growth trend and high citation rates, with a predominant focus on leveraging AI to enhance diagnostic accuracy, particularly in neuro-oncology. Publications were concentrated in specialized, high-impact journals and predominantly originated from authors and institutions in high-income, technologically advanced Northern Hemisphere countries, where scientific collaboration played a foundational role in driving research advancements.

By prioritizing strategic collaboration, translational applications, and inclusive innovation, the field can harness AI’s full potential to advance neurosurgical care, ensuring that emerging technologies benefit diverse patient populations and clinical contexts worldwide.

## Data Availability

The datasets supporting the conclusions of this article are available in the Figshare repository, https://doi.org/10.6084/m9.figshare.28726415

## ABBREVIATIONS

AI: Artificial intelligence
CSV: Comma-separated values
VR: Variation rate
LPI: Lotka’s productivity index
NC: Number of citations
CS: CiteScore
SJR: SCImago Journal Rank
SI: Subramanyan’s index
Ndoc: Number of Documents
N/A: Not assigned.

## DECLARATIONS

### Ethics Approval and Consent to Participate

Not applicable.

### Consent for Publication

Not applicable.

### Competing Interests

The authors declare that they have no competing interests.

### Funding

This research did not receive any specific grant from funding agencies in the public, commercial, or not-for-profit sectors.

### Authors’ Contributions

Conceptualization: Hector Julio Piñera-Castro.

Data curation: Hector Julio Piñera-Castro.

Formal analysis: Hector Julio Piñera-Castro, Christian Borges-García.

Investigation: Hector Julio Piñera-Castro, Christian Borges-García.

Methodology: Hector Julio Piñera-Castro.

Resources: Hector Julio Piñera-Castro.

Supervision: Hector Julio Piñera-Castro.

Visualization: Hector Julio Piñera-Castro.

Writing – original draft: Hector Julio Piñera-Castro, Christian Borges-García.

Writing – review & editing: Hector Julio Piñera-Castro, Christian Borges-García.

## Notes

### Competing Interest Statement

The authors have declared no competing interest.

